# Estimating young adult uptake of smoking by area across the UK

**DOI:** 10.1101/2024.06.25.24309461

**Authors:** Sarah E. Jackson, Harry Tattan-Birch, Nicholas S Hopkinson, Jamie Brown, Lion Shahab, Laura Bunce, Anthony A Laverty, Deborah Arnott

**Affiliations:** Department of Behavioural Science and Health, University College London, London, UK; SPECTRUM Consortium, Edinburgh, UK; National Heart and Lung Institute, Faculty of Medicine, Imperial College London, London, UK; Action on Smoking and Health, UK; School of Public Health, Imperial College London, London, UK

**Author notes:** Corresponding author: Dr Sarah Jackson, Department of Behavioural Science and Health, University College London, 1-19 Torrington Place, London WC1E 7HB, UK.

**Keywords:** smoking, initiation, region, constituency, Great Britain, England, Wales, Scotland, Northern Ireland

## Abstract

**Background:** There is majority support in parliament and across the United Kingdom (UK) to implement a ‘smokefree generation’ policy which would increase the legal age of sale of tobacco from 18 by one year each year from 2027 onwards, such that people born on or after 1 January 2009 could never legally be sold tobacco. To explore the potential impact this policy could have, we estimated the number of young adults (18–25y) currently taking up smoking each year by area across the UK.

**Methods:** Using data from the Office for National Statistics (ONS), Annual Population Survey (APS), and Smoking Toolkit Study (STS), we estimated the total number of 18–25-year-olds in the UK taking up smoking each year, based on national estimates of population size (ONS) and the proportion who reported ever having been a regular smoker (STS). We used local data on adult smoking rates (APS) to apportion the national estimated number of young adults taking up smoking to specific areas.

**Results:** Around 127,500 18–25-year-olds in the UK start smoking regularly each year (349 each day); 105,700 each year (290 each day) in England, 11,500 (32) in Scotland, 6,500 (18) in Wales, and 3,800 (10) in Northern Ireland. Estimates of uptake varied across localities: for example, North East Lincolnshire had the highest proportion of young adults taking up smoking each year (3.96%) and Wokingham had the lowest (0.85%).

**Conclusions:** Despite reductions in smoking prevalence over recent decades, hundreds of young adults in the UK start smoking every day. Data on rates of uptake among individual local authorities can be used to focus attention locally prior to the introduction of new age of sale laws.

## Introduction

Tobacco smoking is uniquely harmful and remains a leading cause of disease, disability, and premature death globally.^1,2^ Two out of every three people who do not manage to quit will die from their smoking.^3^

Most people take up smoking when they are young,^4,5^ underestimating the short-term risks and not expecting it to become a life-long habit.^6^ However, they quickly become addicted^7^ and find it difficult to quit in later life. The majority (54%) of smokers in England want to stop^8^ and three-quarters say they would never have started if they had the choice again.^9^ Starting to smoke at any age has severe health consequences in the long run, but these are particularly pronounced among those who start young.^10^

To reduce the number of young adults taking up smoking and alleviate the burden of smoking-related death and disease for the next generation, the United Kingdom (UK) Government intended to implement a ‘smokefree generation’ policy which would increase the legal age of sale of tobacco from 18 by one year each year from 2027 onwards, such that people born on or after 1 January 2009 would never legally be sold tobacco.^11^ This policy was endorsed by the Chief Medical Officers of all four UK nations (England, Wales, Scotland, and Northern Ireland)^12^ and has widespread public^13^ and parliamentary^14^ support. A bill to enact the legislation was progressing through parliament – and passed a second reading by a majority of 316^14^ – but did not pass into law before a snap election was called in May 2024. Both the party of government and the opposition have committed to bringing the legislation back after the election.

To offer insight into the potential impact this policy could have, this study aimed to estimate the number of young adults (18–25y) currently taking up smoking each year by local area across the UK.

## Methods

### Estimating the number of 18–25-year-olds in the UK taking up smoking

The initial analysis was based on data from the Smoking Toolkit Study, a nationally representative monthly cross-sectional survey of adults (≥16 years) in Great Britain.^15,16^ We analysed data from 7,080 respondents aged 16–25 years surveyed between January 2022 and January 2024.

Smoking status was assessed by asking participants which of the following best applied to them: (a) I smoke cigarettes (including hand-rolled) every day; (b) I smoke cigarettes (including hand-rolled), but not every day; (c) I do not smoke cigarettes at all, but I do smoke tobacco of some kind (e.g., pipe, cigar or shisha); (d) I have stopped smoking completely in the last year; (e) I stopped smoking completely more than a year ago; (f) I have never been a smoker (i.e., smoked for a year or more). Those who responded (a)–(e) were considered ever regular smokers.

Weighted logistic regression was used to model ever regular smoking by age, with age modelled non-linearly using restricted cubic splines (with three knots placed at the 5th, 50th, and 95th percentiles). We used this model to estimate the proportion of 17- and 25-year-olds who have ever regularly smoked, incorporating information from all participants aged 16–25 (rather than just those aged 17 and 25).

We assumed a constant rate of uptake of smoking across each year of ageing, which we calculated as:

> (proportion of 25-year-olds who have ever regularly smoked – proportion of 17-year-olds who have ever regularly smoked) / 8 years = rate of uptake

Using 2021 Office for National Statistics mid-year estimates for population size,^17^ we then estimated the number of 18–25-year-olds in the UK who start smoking regularly each year (assuming the rate of uptake is the same as the Smoking Toolkit Study estimate for Great Britain) as:

> rate of uptake * number of 18–25-year olds = number of 18–25-year-olds taking up smoking each year

For this estimate, we rounded percentages to one decimal place and numbers >1,000 to the nearest 1,000.

### Estimating geographic differences in the number of 18–25-year-olds taking up smoking

This UK estimate was then split across geographical areas according to their adult smoking prevalences, based on the assumption that there was likely to be a greater proportion of young adults taking up smoking in areas that have more adult smokers (i.e., regional differences in smoking uptake would be proportionate to regional differences in adult smoking prevalence).

We obtained data on adult (≥18 years) and young adult (18–25 years) population sizes and adult smoking prevalence in the UK, overall and by nation (England, Scotland, Wales, and Northern Ireland), government office region in England, and upper-tier local authority and unitary authority areas of England, Wales, Scotland, and Northern Ireland. We used the 2021 Office for National Statistics mid-year estimates for population size^17^ and the 2022 Annual Population Survey estimates of smoking prevalence.^18^

For each locality, we calculated the number of adult smokers (local adult population size x local smoking prevalence) and the proportion of UK adult smokers that this represented (local number of adult smokers / total number of adult smokers in the UK). We then apportioned our estimate of the total UK number of new young adult smokers according to the proportion of the total adult smoking population in that locality. We calculated the local number (total number of young adults in the UK taking up smoking each year x local proportion of UK adult smokers) and proportion (local number of young adults taking up smoking each year / local young adult population size) of young adults taking up smoking each year.

The analyses were not pre-registered and should be considered exploratory.

## Results

The Smoking Toolkit Study analysis suggested the prevalence of ever regular smoking increases from 21.9% at 17 years to 37.7% by age 25. Assuming a constant rate of uptake, this equates to 2.0% uptake across any year of aging between ages 18 to 25 ((37.7%-21.9%) / 8 years). There are ∼6,375,000 people aged 18–25 in the UK.^17^ We therefore estimated that 127,500 (6,375,000 * 2.0%) 18–25-year-olds in the UK start smoking regularly each year.

The estimated proportion of 18–25-year-olds who take up smoking each year is presented as a heat map by local authority and unitary authority areas across the UK in **Figure 1**. Tables with daily, weekly, monthly, and annual figures for each UK nation and region in England are provided in

**Figure 1.**
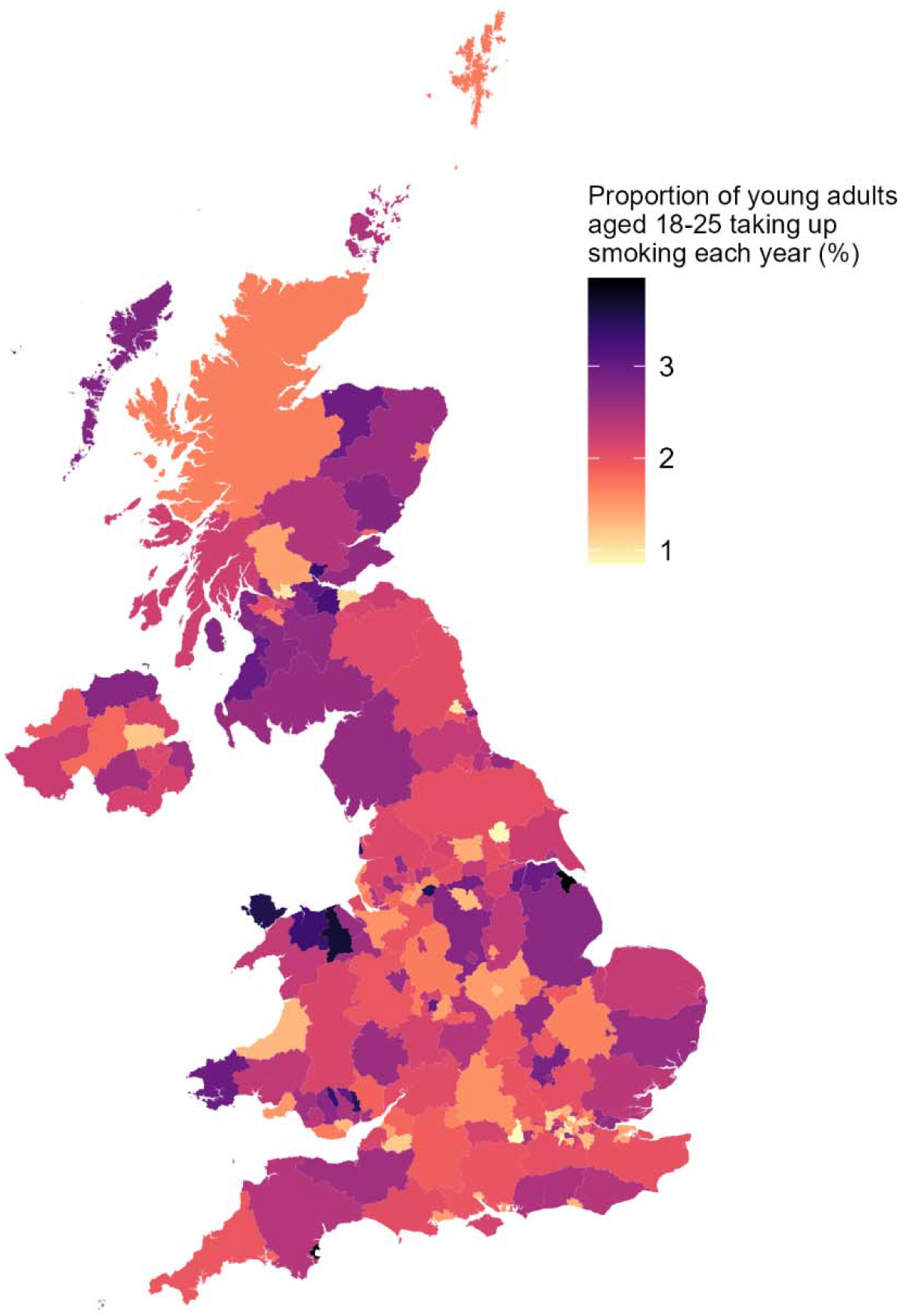
Estimated proportion of young adults aged 18–25 who start smoking in the UK each year. For each locality, data shown are the estimated total number of young adults taking up smoking in the UK each year apportioned according to the proportion of the total adult smoking population in that locality, divided by the local young adult population size.

**Table 1**; corresponding estimates for local authority and unitary authority areas are available in the Supplementary File.

**Table 1.** Estimates of smoking uptake among 18**–**25-year-olds by UK nation and region in England

|  | Adult population<sup>1</sup> | Smoking prevalence, %<sup>2</sup> | No. of adult smokers<sup>3</sup> | % of UK smokers | Young adult population (18–25y)<sup>1</sup> | Number of new smokers<sup>4</sup> per... |  |  |  |
| --- | --- | --- | --- | --- | --- | --- | --- | --- | --- |
|  |  |  |  |  |  | Year | Month | Week | Day |
| United Kingdom | 53,188,000 | 12.9 | 6,861,300 | 100.0 | 6,375,000 | 127,500 | 10,625 | 2,452 | 349 |
| Nation |  |  |  |  |  |  |  |  |  |
| England | 44,775,000 | 12.7 | 5,686,400 | 82.9 | 5,392,000 | 105,668 | 8,806 | 2,032 | 290 |
| Scotland | 4,455,000 | 13.9 | 619,200 | 9.0 | 516,000 | 11,507 | 959 | 221 | 32 |
| Wales | 2,489,000 | 14.1 | 350,900 | 5.1 | 291,000 | 6,521 | 543 | 125 | 18 |
| Northern Ireland | 1,470,000 | 14.0 | 205,700 | 3.0 | 177,000 | 3,823 | 319 | 74 | 10 |
| Region in England |  |  |  |  |  |  |  |  |  |
| East Midlands | 3,890,000 | 14.0 | 544,600 | 7.9 | 480,000 | 10,120 | 843 | 195 | 28 |
| East England | 5,018,000 | 13.2 | 662,300 | 9.7 | 546,000 | 12,308 | 1,026 | 237 | 34 |
| London | 6,904,000 | 11.7 | 807,800 | 11.8 | 925,000 | 15,011 | 1,251 | 289 | 41 |
| North East | 2,122,000 | 13.1 | 278,000 | 4.1 | 255,000 | 5,165 | 430 | 99 | 14 |
| North West | 5,860,000 | 13.4 | 785,300 | 11.5 | 715,000 | 14,593 | 1,216 | 281 | 40 |
| South East | 7,358,000 | 11.5 | 846,100 | 12.3 | 826,000 | 15,724 | 1,310 | 302 | 43 |
| South West | 4,626,000 | 11.9 | 550,500 | 8.0 | 526,000 | 10,229 | 852 | 197 | 28 |
| West Midlands | 4,663,000 | 13.4 | 624,800 | 9.1 | 576,000 | 11,610 | 968 | 223 | 32 |
| Yorkshire and the Humber | 4,335,000 | 13.1 | 567,900 | 8.3 | 543,000 | 10,553 | 879 | 203 | 29 |

<sup>1</sup> Office for National Statistics 2021 mid-year population estimates.<sup>2</sup> Annual Population Survey, 2022.<sup>3</sup> Adult population x smoking prevalence.<sup>4</sup> Estimated number of 18–25-year-olds taking up smoking.
Note: estimated numbers of new smokers by UK nation or region in England do not sum precisely to totals due to rounding.

Of the 6.375 million 18–25-year-olds across the UK, an estimated 349 start to smoke each day: 290 in England, 32 in Scotland, 18 in Wales, and 10 in Northern Ireland. There was substantial variation in the proportion of young adults taking up smoking across localities. North East Lincolnshire and Torbay had the highest rates of uptake (3.96% and 3.90% annually): each month, 42 from a population of 13,000 in North East Lincolnshire and 33 from a population of 10,000 in Torbay started to smoke regularly. Uptake was lowest in Wokingham and York (0.85% and 0.87% annually), with 10 and 22 young adults starting to smoke regularly each month from populations of 14,000 and 31,000, respectively.

## Conclusions and limitations

This analysis shows that despite reductions in smoking prevalence over recent decades,^18^ hundreds of young adults in the UK start smoking every day. Raising the legal age of sale is likely to be an effective measure to reduce this, based on evidence that increases from 16 to 18 in the UK in 2007^19–21^ and increases to 21 in the US^22–26^ were associated with decreases in smoking among age groups that could not legally able to buy tobacco. Steadily increasing the legal age of sale further will likely reduce the chance of people becoming addicted at any point in life, given older age of smoking initiation is associated with lower levels of lifetime nicotine dependence.^27^ As well as the overall scale of uptake, the data here on rates of uptake among individual local authorities can be used to focus attention locally on the value to their communities of the national policy to raise the age of sale to help prevent smoking initiation at any age.

It is important to acknowledge that this type of estimation has limitations. The data presented are extrapolated from survey data, so are necessarily approximate. For some local and unitary authority areas, Annual Population Survey estimates of smoking prevalence were based on small samples so may be unreliable (for reference, sample sizes are provided in the Supplementary File). The Smoking Toolkit Study does not cover Northern Ireland, so the rate of uptake was estimated based on data collected in Great Britain, rather than the whole of the UK. We also assumed that current population sizes were similar to 2021 and that local variation in rates of smoking uptake would correspond to variation in adult smoking rates.

## Data Availability

Data are available on the Open Science Framework (https://osf.io/nu2rp/)

## Declarations

### Ethics approval

Ethical approval for the STS was granted originally by the UCL Ethics Committee (ID 0498/001). The data are not collected by UCL and are anonymised when received by UCL.

